# Dose-dependent degeneration of non-cancerous brain tissue in post-radiotherapy patients: A diffusion tensor imaging study

**DOI:** 10.1101/19005157

**Authors:** Szabolcs David, Hamed Y. Mesri, Victor A. Bodiut, Steven H. J. Nagtegaal, Hesham Elhalawani, Alberto de Luca, Marielle E. P. Philippens, Max A. Viergever, Abdallah S. R. Mohamed, Yao Ding, Caroline Chung, Clifton D. Fuller, Joost J. C. Verhoeff, Alexander Leemans

## Abstract

**Background and purpose:** Radiation-induced changes in brain tissue may relate to post-radiotherapy (RT) cognitive decline. Our aim is to investigate changes of the brain microstructural properties after exposure to radiation during clinical protocols of RT using diffusion MRI (dMRI).

**Methods and Materials:** The susceptibility of tissue changes to radiation was investigated in a clinically heterogenic cohort (age, pathology, tumor location, type of surgery) consisting of 121 scans of 18 patients (10 females). The imaging dataset included 18 planning CTs and 103 dMRI scans (range 2-14, median = 6 per patient) assessing pre-operative, post-operative pre-RT and post-RT states. Diffusion tensor imaging (DTI) metrics were estimated from all scans for a region-of-interest based linear relation analysis between mean dose and change in DTI metrics, while partial volume effects were regressed out.

**Results:** The largest regional dose dependency with mean diffusivity appear in the white matter of the frontal pole in the left hemisphere by an increase of 2.61 %/(Gy x year). Full brain-wise, pooled results for white matter show fractional anisotropy to decrease by 0.85 %/(30Gy x year); mean diffusivity increase by 9.17 %/(30Gy x year); axial diffusivity increase by 7.30%/(30Gy x year) and radial diffusivity increases by 10.63%/(30Gy x year).

**Conclusions:** White matter is susceptible to radiation with some regional variability where diffusivity metrics demonstrate the largest relative sensitivity. This suggests that dMRI is a promising tool in assessing microstructural changes after RT, which can help in understanding treatment-induced cognitive decline.

## 1 Introduction

Radiation therapy (RT) is a common treatment procedure for both primary and metastatic tumors in the brain, often in combination with surgery and chemotherapy. As radiation impact is not selective to tumor cells, the efficacy of RT is hindered by the radiosensitivity of healthy surrounding tissue [1,2]. The effect of radiation on brain tissue is dynamic and involves structures outside the targeted tumor volume, directly or indirectly [3,4]. Despite the substantial advances in RT technology and application [5,6], the regional differences in sensitivity to radiation dosing is not well-documented, which is especially true for white matter (WM) structures. Current clinical protocols include guidelines for maximum dose for brain parenchyma and several organs at risk for which the direct structure-function relationship is well-understood, or are visible on CT scans, e.g.: brain stem, optic chiasm, optic nerves, cochlea and hippocampi [7]. However, regional constraints on WM and other structures are not part of the standard consideration.

Diffusion MRI (dMRI) is the preferred non-invasive imaging technique to study WM structure in the brain [8–10]. The fundamental principle behind dMRI is that the Brownian motion of water molecules is dependent on the surrounding tissue structure [11]. WM is a highly structured tissue type, which introduces diffusion anisotropy along a certain direction and restricts the extent of local diffusion. This is reflected in the diffusion tensor imaging (DTI) metrics, such as fractional anisotropy (FA) and mean, radial, and axial diffusivity (MD, RD, and AD, respectively), making dMRI a quantitative method to examine WM anatomy and its microstructural properties. Neuro-oncological research has placed dMRI as a potential technique for tumor diagnosis [12], surgical planning [13], pre-treatment prediction of tumor response [14], monitoring early efficacy of treatment [15] and early WM damage post-radiation [16], and, more recently, late-delayed effects of RT on WM [17].

Radiation-induced WM damage has been reported to include axonal injury, demyelination, neuro-inflammation, and necrosis among others [18–20]. Importantly, these structural deficits seem to correlate in time with both verbal and non-verbal cognitive impairment, including executive functioning, working memory, visuospatial processing, and decision-making [21]. In this context, radiation-induced cognitive impairment has been divided into multiple phases post-RT: acute (<2 weeks), early-delayed (2 weeks to 3 months) and late-delayed (6+ months). Notably, while acute and early-delayed symptoms and damage are mostly temporary, late-delayed damage is considered permanent [22–24]. The introduced cognitive impairment currently occurs in 50-90% of survivors [25,26], and this population is increasing with RT advancement [27,28]. The progressive decline affects the physical and mental health of long-term survivors and impairs the patients’ quality of life (QoL) [22,23]. Therefore, a better understanding of the mechanism of post-irradiation cognitive decline is necessary. High RT doses (>60Gy) are well-documented to cause permanent damage, but emerging evidence shows that even at lower doses (<20Gy) late-delayed damage can still occur [29]. Information on regional WM sensitivity to RT dosage in the long term is crucial for better RT planning and ensuring optimal QoL after treatment.

In this work, we aim to quantify the long-term or late-delayed effects of RT on brain tissue microstructures using dMRI. Specifically, we are interested in how the susceptibility of different structures to radiation varies across regions and dose levels.

## 2 Materials and Methods

### 2.1 Cohort

We retrospectively identified patients who were treated with RT at the Department of Radiation Oncology MD Anderson Cancer Center, University of Texas, Houston, Texas, USA. Patients were eligible for inclusion upon met the following criteria: inclusion of a pre-RT MRI scan, which is defined as the baseline (BL) of a scan within one month of the RT start, and follow-up (FU) scans between 3 and 24 months to exclude the acute-early and very late effects. The selection resulted in 18 patients (10 females). All patients received intensity-modulated RT (IMRT); most of them were treated to 60 Gy in 30 fractions, others were recalculated to a total equivalent dose in standard 2 Gy fractionation (EQD2) using biologically equivalent dose principles using the linear quadratic model with an α/β ratio of 2 Gy [30]. Detailed information of treatment and demographic factors are shown in Table 1. This study was approved by the institutional review board and informed consent was obtained from all subjects.

**Table 1.** Demographics and tumor characteristics of the patients included in the cohort.

| Patient info | Number of patients |
| --- | --- |
| Sex |  |
| Male | 8 |
| Female | 10 |
| Median age range (years) | 56 (37-76) |
| Pathology (tumor type) |  |
| Anaplastic astrocytoma | 2 |
| Glioblastoma | 13 |
| Gliosarcoma | 1 |
| Oligodendroglioma | 1 |
| Progressive anaplastic oligodendroglioma | 1 |
| Tumor location |  |
| Brain stem | 1 |
| Left frontal | 5 |
| Left frontoparietal | 1 |
| Left parietal | 1 |
| Left temporal | 2 |
| Right frontal | 4 |
| Right parietal | 1 |
| Right temporal | 3 |
| Surgery |  |
| Total resection | 8 |
| Subtotal resection | 5 |
| Craniotomy and resection | 3 |
| Partial resection | 1 |
| None | 1 |
| RT dose, Gy (fraction size) |  |
| 60 (2) | 14 |
| 57 (1.9) | 2 |
| 54 (1.8) | 1 |
| 50 (1.85) | 1 |
| Chemotherapy |  |
| Temodar | 17 |
| none | 1 |

### 2.2 Image acquisition

#### 2.2.1 CT acquisition

The planning CT scans were acquired on a Philips Brilliance Big Bore scanner, with a tube potential of 120 kVp, matrix size of 512 × 512 × 87 and 0.98 × 0.98 × 3.0 mm^3^ voxel size.

#### 2.2.2 MRI acquisition

MR images were acquired at multiple time points for each patient on various GE Medical Systems MRI scanners. The dataset included 103 dMRI scans (range 2-14, median = 6 per patient) acquired using 27 gradient directions with a diffusion weighting of b = 1200 s/mm^2^ and one b = 0 s/mm^2^. Other acquisition settings showed large heterogeneity between patients, but within each patient, the images were acquired on the same type of scanner. At 1.5T, examinations were performed with Signa Excite (in 6% of the 103 scans) and Signa HDxt (51%) scanners. For all 1.5T scans, the dMRI acquisition details were: TR = 10 s, TE = 102 ms, voxel size: 0.86 mm × 0.86 mm × 6.5 mm in 58% of all scans. At 3T, the following scanners were employed: Signa Excite <1%, Signa HDx, 2%, Signa HDxt scanner 39%. For all 3T scans, the dMRI acquisition details were: TR = 8.75 s, TE = 91 ms, voxel size: 0.86 mm × 0.86 mm × 3.5 mm in 42% of all scans.

### 2.3 Image processing

All non-dMRI data were processed with in-house algorithms developed in Matlab (Mathworks, Natick, Massachusetts), whereas all dMRI data were processed with *ExploreDTI* [31].

First, the CT images were cropped to increase registration performance and to reduce computational load in later stages. This step resulted in a bounding box around the skull, excluding the neck and the shoulders of the patients. The same cropping was applied for the dose, organs-at-risk (OaR) and planning, clinical, and gross target volume (TV) maps per patient.

dMRIs were corrected for subject motion and eddy current distortions using the non-diffusion-weighted image as the reference, whereupon all images were rigidly registered to the cropped CT image [32]. In order to reduce the blurring effect of multiple registration procedures, the two transformations were concatenated into a single resampling step. During motion correction and subsequently during registration to the CT, the DW gradients were adjusted with the rotational element of the registrations [33].

Robust estimation of the diffusion tensor model was performed with REKINDLE [34]. The region-of-interest (ROI) definition for the analysis was performed by non-linearly registering the ‘wmparc’ brain template and the associated atlas to every individual DWI scan [35], which were already motion-corrected and co-registered to the CT scan. The mentioned atlas contains 179 labels covering the whole brain and is freely available in *Freesurfer* [36]. A graphical overview of the automated image processing pipeline is shown in Fig. 1.

**Fig. 1.**
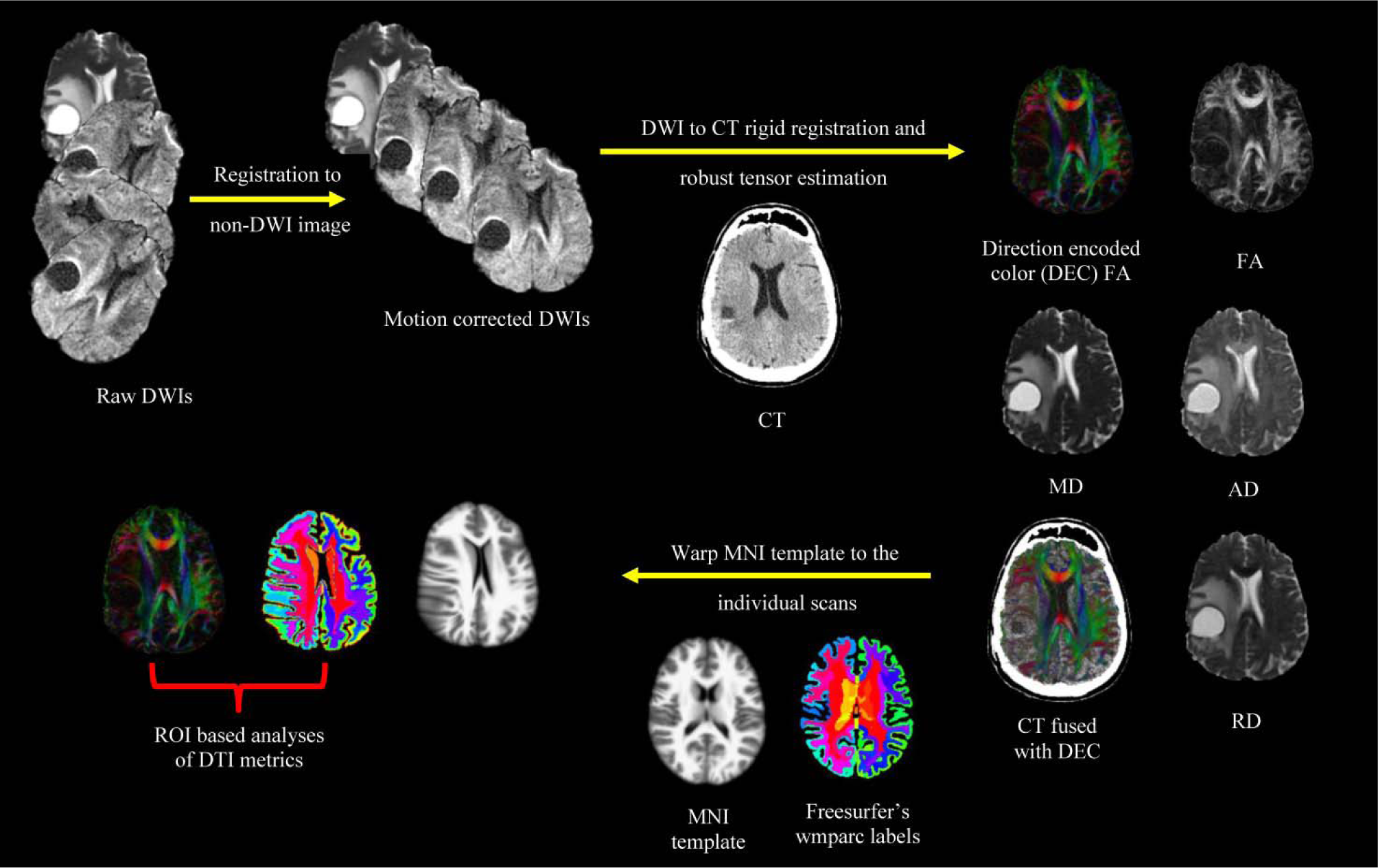
Graphical overview of the image processing pipeline. First, the raw diffusion weighted images (DWIs) registered to the very first non-DW image. Next, all images were further registered to the CT scan followed by a robust diffusion tensor estimation, which leads to the calculation of diffusion tensor imaging scalars as fractional anisotropy (FA), mean-radial-axial diffusivity (MD, RD, and AD, respectively). A standard template from the Montreal Neurological Institute (MNI) and associated labels from *Freesurfer* were non-linearly registered to the already CT-coregistered DWIs. The registered labels allow an atlas-defined ROI based analyses of DTI metrics.

The processing pipeline generates DTI metrics based on atlas-defined ROIs in the native CT space, yielding high spatial alignment between anatomical and DTI data for each timepoint. Finally, equivalent dose in 2 Gy per fraction (EQD2) dose values and DTI metrics from multiple time points were accessible per ROI per patient. We computed the following DTI metrics per ROI: average of FA, MD, AD, RD; volume in mm^3^ and mean of the EQD2 dose. Examples of the CT-dMRI alignment are shown in Figs. 2/a-c, while Figs. 2/d-f show changes of the tissue and the tumor cavity after the start of RT with an exemplar 3D rendering of the inferior temporal white matter in the non-affected hemisphere.

**Fig. 2.**
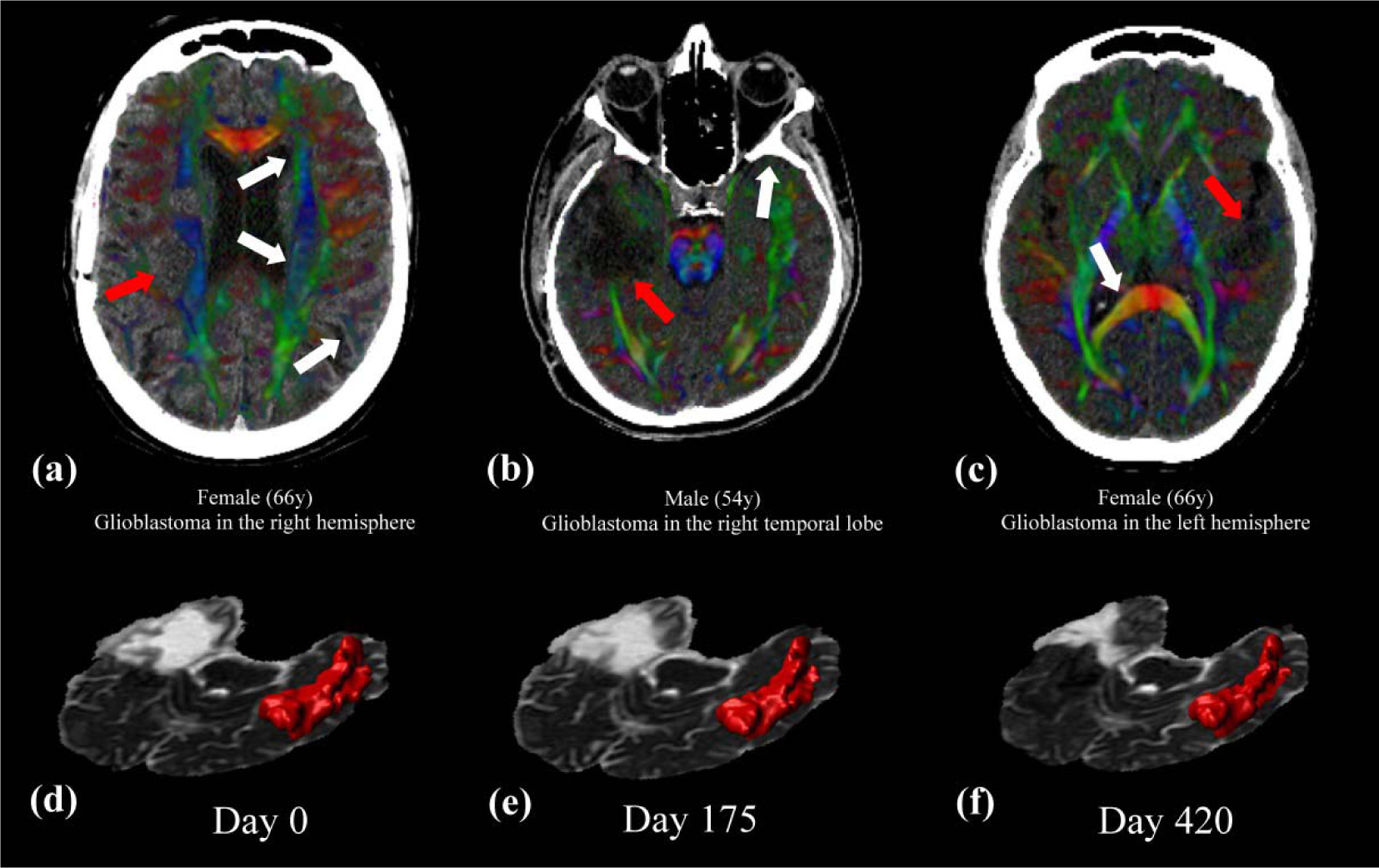
Graphical overview image processing results. Panels a-b-c shows the CT and dMRI alignment via fused CT - direction encoded color (DEC) maps in representative patients, where the background image is the planning CT and the colors show the locally dominant direction of diffusion via the DTI model. The intensity of the color was weighted with fractional anisotropy (FA), which suppresses the color in CSF, because FA is close to 0 in homogeneous media. White arrows pinpoint critical regions of alignment at (a) edge of the ventricles and gyrus, (b) interface of the brain-skull, and at (c) ventricle – white matter interface. Red arrows show the tumor cavity, where the absence of strong anisotropic diffusion resulted in no color coding. In the bottom row: (d)-(e)-(f) show the changes of the tumor cavity of patient (b) with mean diffusivity (MD) as the background image. An example of an atlas region is shown in red: the 3D rendered region of the inferior temporal white matter in the non-affected hemisphere.

To avoid data contamination from tumor cavities, remaining cancerous tissue, or surgical scars, all ROIs that overlapped the GTV were excluded from the analysis. Also, ROIs covering the ventricles and cerebrospinal fluid CSF were ignored. Moreover, regions with the atlas label ‘Unsegmented White Matter’ were excluded due to its undefined nature. In total, 16 of the original 179 labels were ignored and a total of 6.85% of the 18*163 regions were not included for further analysis.

### 2.4 Longitudinal analysis

Analyzing data from unequally time-spaced acquisitions can be challenging, given that standard statistical models of repeated measures are not applicable, like paired t-tests or ANOVAs. One solution is to perform regression analysis for every ROI in every patient with the following details: the timing of the follow-up scans is the independent variable, while the mean FA, MD, AD and RD per ROI are the dependent variables. The pre-RT MRI scan is defined as the baseline (BL) with day = 0. This calculation yields in the rate of change of the DTI metrics. In another pipeline, we included the volume of the ROIs as a covariate of no-interest as Vos et al. showed that partial voluming can act as a hidden covariate in DTI analyses [37]. Calculations described above were performed with *FSL* utility tool *fsl_glm* [38].

Next, the estimated coefficients from all four dMRI metrics were correlated with the mean EQD2 dose per ROI using a permutation test with 5000 iterations performed with the permutation analysis of linear models (PALM) toolbox version alpha104, a Matlab based open-source software package [39–41]. We used nonparametric permutations as they proved efficient in eliminating false positive results when compared with parametric methods [42]. Significance of a correlation was determined at p_corr_ < 0.05 using family-wise error rate (FWER) adjustment to correct for multiple comparisons. Tail approximation was used for faster calculations [43]. Age at the time of the diagnosis and sex of the patients were included as nuisance regressors. An effect size measure, the Cohen’s D, is reported to grade the practical significance of the results [44]. A graphical overview of the longitudinal analysis is shown in Fig. 3.

**Fig. 3.**
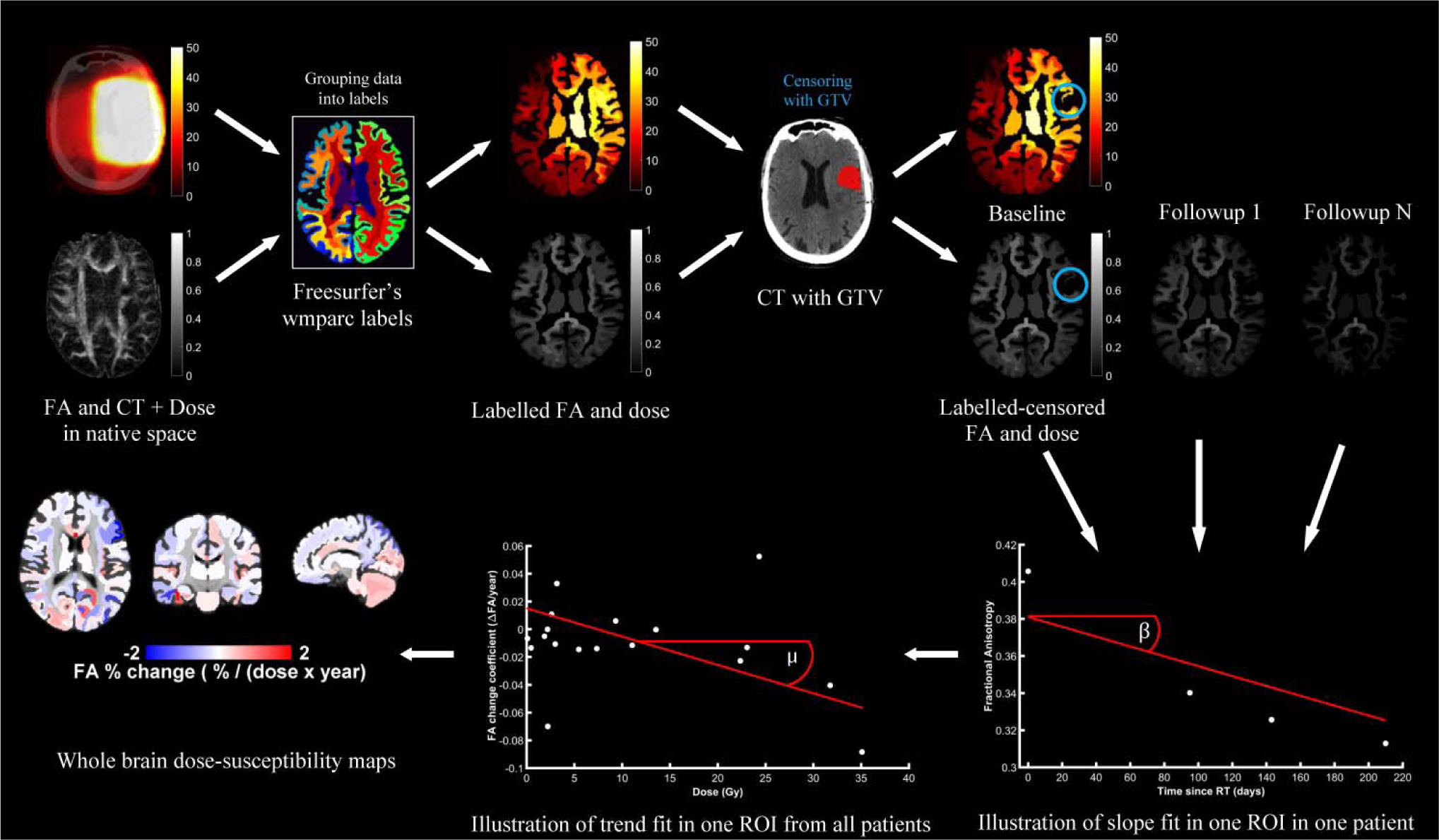
Graphical overview of the longitudinal analysis. First the motion-corrected dMRI metrics are grouped by atlas labels as well as doses. Regions that overlapped with the GTV were excluded from the analysis. The rate of metric change after RT was calculated for every region in every patient. The individual rates were then pooled to calculate the dose dependency of the metric changes. Repeating the procedure for all regions and metrics leads to whole brain dose-susceptibility maps.

Additionally, variance groups (VGs) were defined based on the number of scans per patient, since permutations are only valid within the same VG. Defining the VGs is needed because the variance of the changes in the metrics is not equal in all subjects and depends on the number of scans used. This introduces heteroscedasticity in the metrics, which we accounted for by limiting permutations via VGs.

## 3 Results

Fig. 4 shows the whole-brain dose susceptibility map as measured with MD for those regions which were statistically significant using a critical p-value = 0.05 as significance threshold. In the majority of brain regions, MD increases with increasing dose. Also, some regional differences appear, with the largest dose dependencies in the anterior part of the Corpus Callosum by 1.77 % / (Gy x year); in the caudal anterior cingulate cortex (ACC) in the right hemisphere (also known as Brodmann areas 24, 32 and 33) by 1.78 % / (Gy x year); and in the white matter of the frontal pole (also known as Brodmann area 10) in the left hemisphere by 2.61 % / (Gy x year). Supplementary Figs. 1, 2 and 3 show the spatial distribution of dose susceptibility maps of FA, AD, and RD, respectively. Supplementary Fig. 4 shows the overall mean dose distribution, where mostly the deep grey matter (GM) regions received the highest dose on average. Supplementary Fig. 5, 6, 7, 8 and 9 shows the coefficients of change along with the 95% confidence interval for regions in the left hemisphere of the cortex; right hemisphere of the cortex; left hemisphere of the white matter; right hemisphere of the white matter; and the deep GM regions, along with the brainstem, segments of the corpus callosum and the cerebellum; respectively.

**Fig. 4.**
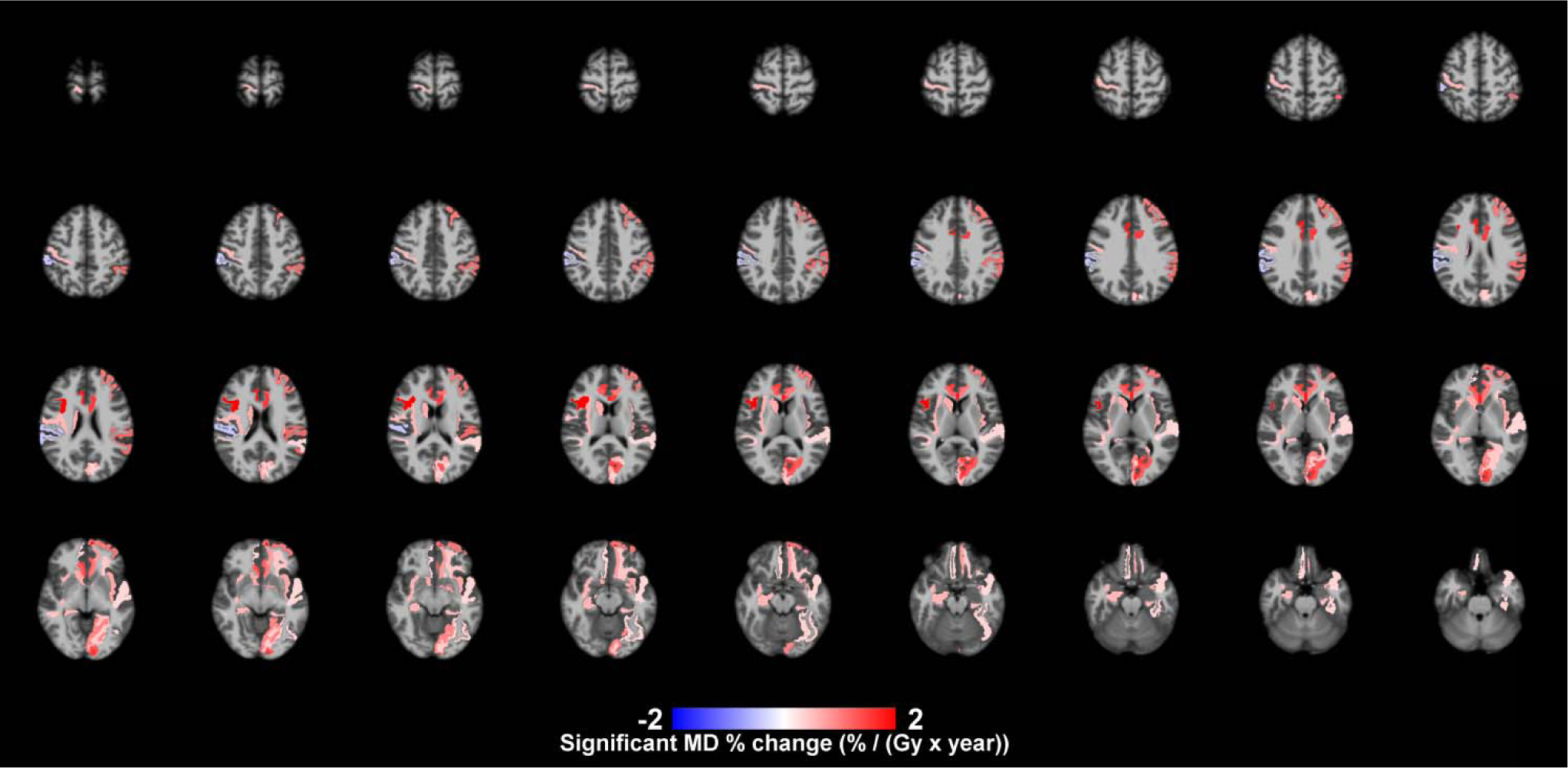
Whole-brain dose-susceptibility map measured with MD, after significance thresholding. The color shading represents the effect size in terms of percentage change in a year per unit dose, while the background image is the MNI template (radiological view: left on the image is right in the brain and vice versa).

Table 2. shows pooled results for 30 Gy per cortical GM, deep GM, and WM; with and without considering region volume as a covariate of no-interest. After accounting for volume, all metrics increased, which resulted in elevated diffusivity susceptibilities and a diminished FA-dose relation. Supplementary Table 1 shows the cohort-based mean values cortical GM, deep GM, and WM. In WM, FA shows hardly any response to the applied dose, whereas the increase in diffusivities shows larger susceptibility to dose than FA in all tissue types. The increase of RD in WM is nearly 50% greater than AD, meaning that the diffusivity increase perpendicular to the dominant fiber direction is much larger than the increase along the dominant direction for a given dose. In cortical GM, RD is nearly 15% higher than AD, whereas in deep GM, RD is 10% lower than AD.

**Table 2.** Pooled percentage rates of tissue degradation with different DTI metrics.

|  | With ROI volume as nuisance |  |  |  | Without ROI volume as nuisance |  |  |  |
| --- | --- | --- | --- | --- | --- | --- | --- | --- |
|  | Change (% / (30 Gy x year) ) |  |  |  | Change (% / (30 Gy x year) ) |  |  |  |
| DTI metric | FA | MD | AD | RD | FA | MD | AD | RD |
| Cortical GM | -2.31 | 8.55 | 7.81 | 8.96 | -3.82 | 7.31 | 6.26 | 8.04 |
| Deep GM | 2.43 | 7.92 | 8.07 | 7.41 | -3.79 | 6.46 | 5.54 | 6.81 |
| WM | -0.85 | 9.17 | 7.30 | 10.63 | -2.39 | 8.22 | 6.18 | 9.73 |

## 4 Discussion

In this work, we presented a systematic ROI-based analysis of microstructural properties of the brain in patients after cranial RT. The results show that the changes estimated with DTI are tissue-dependent and are much more pronounced in the diffusivity metrics (MD, AD and RD) than in FA. Studies on tissue-dose interaction, as the current one, aid in understanding radiation-induced cognitive decline, which may be the consequence of, but not restricted to, demyelination and loss of axonal integrity with increasing radiation doses. Research in neuroscience, especially on aging [47,48] and cognitive impairment [49,50] have already established a link between WM properties as determined with DTI and mental health of the participant [45]. Information about regional sensitivity to RT is the first step towards new cognition-sparing radiation treatment planning resulting in lower adverse side effects.

Volume changes in brain structures have been well-studied because of their potential link to cognitive functions, such as the volume-memory relationship in the hippocampus [46–49]. RT-related changes in hippocampal volume seem to be consistent, show dose-dependency, and correlate with follow-up memory assessments, thereby promoting the hippocampal volume as an important biomarker and an organ to protect from radiation [23,45,50–53]. In summary, region volume is affected by RT and for this reason we incorporated volume as a nuisance during statistical inference. This led to increase of all metric change rates and resulted in having the largest effect on FA. It confirms the findings of Vos et al [54] that the volume of a bundle or region can act as a hidden covariate in quantitative analyses. Furthermore, the effects of partial voluming on FA are remarkable; neglecting it would lead to a conceptually different outcome. The partial volume effects (PVE) are reduced in the DTI metric changes, therefore the results represent only the microstructural-related changes as estimated with DTI. Studies neglecting PVEs are prone to unreliable analysis. To the best of our knowledge, previous works on regional tissue sensitivity to RT discuss volumes only within ROIs receiving certain dose levels or aim to avoid PVE by restricting the masking to connected voxels that only include WM structure by excluding voxels with FA lower than 0.2 [17,29,55].

Previous studies investigating tissue properties with dMRI and radiation are in line with our work: an overall decrease of FA and an increase of the various diffusivities [56–59]. The exact underlying mechanisms are ambiguous, since changes of FA are not specific to one well-defined procedure, like demyelination, scarification [60] and axonal loss [61]. However, some relations are possible to recover. The appearance of edema reduces FA while it increases MD [62,63], which is a result of increased presence of free water [64]. With the measurements of AD and RD, it is possible to look into the change of diffusivities in depth. RD is 50% more sensitive to radiation than AD in WM, meaning that diffusion becomes increasingly isotropic with a progressive dose. These finding are consistent with previous studies, confirming that the increase in RD overcomes the increase in AD [19,55]. Similarly, an increase in RD has been suggested to relate to demyelination in human [65] and animal [20] RT studies. It is important to note that, while cortical and deep GM show large relative changes in FA, these changes are negligible in absolute terms since FA in GM is close to 0 due to the absence of anisotropic media at the typical acquisition resolution.

Regional differences appear, but it remains doubtful whether these are genuine or artefactual as previous examples pooled the bilateral regions into a single one, forcing symmetrical results [55]. Clearly, the environment for testing tissue susceptibility to dose is not equal for all regions as dose distribution is non-homogeneous. Much larger clinical cohorts are necessary in the future to selectively balance out this tendency, which originates from the spatial heterogeneity of brain tumors [66–68].

Tissue changes after radiotherapy can be also addressed via analyzing T1-weighted (T1w) images. In this work, however, we excluded the T1w scans from the analysis because of highly anisotropic voxel sizes. The larger slice thickness will cause significant PVEs that will impede a meaningful morphometric analysis. New guidelines [69] have been proposed to set up robust acquisition protocols and processing pipelines to tackle volumetric [70] and surface features (e.g., cortical thickness [71]). T1-based brain analysis has a long history in neuroscience, revealing similar changes to RT as in healthy aging, but in a period of decades as opposed to our maximum 24 months long follow-up period [72]. Accelerated aging as a consequence of RT has been also described by non-MRI studies investigating physiologic frailty [73] and accelerated neurocognitive function decline [74].

For the modeling of tissue changes we assumed a linear relationship between the tissue characteristics and time passed since RT. We further assumed that tissue change relates linearly to the applied dose. While this framework is widely applied by the RT-community [17,29,55,75], these assumptions may only approximate the true underlying relations. Further biophysical modeling is necessary to establish a more accurate representation of the tissue-dose relation and, in turn, to provide a better understanding of the underlying tissue breakdown mechanisms [76].

In this study, we considered the relation of DTI metrics and radiation, which - regardless of the significance of the association - does not necessarily translate into clinically significant effects. Furthermore, it provides no information of the disturbed functionality of the regions, as one may consider that a certain change in, for instance, the hippocampi can have different consequences than the same change in the temporal cortex.

The current study uses DTI as a means of diffusion modeling. However, it has been implied that DTI has some critical limitations, like the inability to resolve crossing fibers [77,78]. Recent developments in diffusion MRI suggest [79] that more advanced models than DTI, like diffusional kurtosis imaging [80–82] or spherical deconvolution [83–85], should be taken into consideration.

## 5 Conclusion

In this work, we investigated radiation-induced changes in the brain measured with DTI in a clinical population. Our findings suggest that radiation-caused damage of brain tissue follows a similar process as aging, but in a much faster fashion. Future studies should incorporate auxiliary modalities as functional MRI or perfusion mapping [86] as well as behavioral and cognitive tests for a more detailed investigations to reveal the causal relationship between changed tissue characteristics and changes in mental health.

## Data Availability

The data generated during the study is available from the corresponding author on reasonable request by a qualified individual or third-party, subject to authorization of relevant regulatory bodies.

## References

[1] Dawson LA, Jaffray DA. Advances in image-guided radiation therapy. J Clin Oncol 2007;25:938–46. doi:10.1200/JCO.2006.09.9515.

[2] Kelley K, Knisely J, Symons M, Ruggieri R. Radioresistance of brain tumors. Cancers (Basel) 2016;8:1–28. doi:10.3390/cancers8040042.

[3] Schultheiss TE, Kun LE, Ang KK, Stephens LC. Radiation response of the central nervous system. Int J Radiat Oncol Biol Phys 1995;31:1093–112. doi:10.1016/0360-3016(94)00655-5.

[4] Tofilon PJ, Fike JR. The radioresponse of the central nervous system: a dynamic process. Radiat Res 2000;153:357–70.

[5] Baskar R, Lee KA, Yeo R, Yeoh KW. Cancer and radiation therapy: Current advances and future directions. Int J Med Sci 2012;9:193–9. doi:10.7150/ijms.3635.

[6] Lagendijk JJWW, Raaymakers BW, Raaijmakers AJEE, Overweg J, Brown KJ, Kerkhof EM, et al. MRI/linac integration. Radiother Oncol 2008;86:25–9. doi:10.1016/j.radonc.2007.10.034.

[7] Scoccianti S, Detti B, Gadda D, Greto D, Furfaro I, Meacci F, et al. Organs at risk in the brain and their dose-constraints in adults and in children: A radiation oncologist’s guide for delineation in everyday practice. Radiother Oncol 2015;114:230–8. doi:10.1016/j.radonc.2015.01.016.

[8] Jeurissen B, Descoteaux M, Mori S, Leemans A. Diffusion MRI fiber tractography of the brain. NMR Biomed 2019;32:e3785. doi:10.1002/nbm.3785.

[9] Assaf Y, Johansen-Berg H, Thiebaut de Schotten M. The role of diffusion MRI in neuroscience. NMR Biomed 2019;32:e3762. doi:10.1002/nbm.3762.

[10] Lebel C, Treit S, Beaulieu C. A review of diffusion MRI of typical white matter development from early childhood to young adulthood. NMR Biomed 2019;32:e3778. doi:10.1002/nbm.3778.

[11] Kiselev VG. Fundamentals of diffusion MRI physics. NMR Biomed 2017;30:1–18. doi:10.1002/nbm.3602.

[12] Kono K, Inoue Y, Nakayama K, Shakudo M, Morino M, Ohata K, et al. The role of diffusion-weighted imaging in patients with brain tumors. AJNR Am J Neuroradiol 2001;22:1081–8.

[13] Nimsky C, Ganslandt O, Hastreiter P, Wang R, Benner T, Sorensen AG, et al. Intraoperative Diffusion-Tensor MR Imaging: Shifting of White Matter Tracts during Neurosurgical Procedures—Initial Experience. Radiology 2005;234:218–25. doi:10.1148/radiol.2341031984.

[14] Mardor Y, Roth Y, Ocherashvilli A, Spiegelmann R, Tichler T, Daniels D, et al. Pretreatment Prediction of Brain Tumors Response to Radiation Therapy Using High b-Value Diffusion-Weighted MRI. Neoplasia 2004;6:136–42. doi:10.1593/neo.03349.

[15] Chenevert TL. Diffusion Magnetic Resonance Imaging: an Early Surrogate Marker of Therapeutic Efficacy in Brain Tumors. J Natl Cancer Inst 2000;92:2029–36. doi:10.1093/jnci/92.24.2029.

[16] Price SJ, Burnet NG, Donovan T, Green HAL, Peña A, Antoun NM, et al. Diffusion tensor imaging of brain tumours at 3 T: A potential tool for assessing white matter tract invasion? Clin Radiol 2003;58:455–62. doi:10.1016/S0009-9260(03)00115-6.

[17] Zhu T, Chapman CH, Tsien C, Kim M, Spratt DE, Lawrence TS, et al. Effect of the Maximum Dose on White Matter Fiber Bundles Using Longitudinal Diffusion Tensor Imaging. Int J Radiat Oncol Biol Phys 2016;96:696–705. doi:10.1016/j.ijrobp.2016.07.010.

[18] Leeds NE, Fuller GN, Tassel P Van, Maor MH, Sawaya RE, Levin VA, et al. Malignant Gliomas: MR Imaging Spectrum of Radiation Therapy- and Chemotherapy-induced Necrosis of the Brain after Treatment. Radiology 2000;217:377–84. doi:10.1148/radiology.217.2.r00nv36377.

[19] Nagesh V, Tsien CI, Chenevert TL, Ross BD, Lawrence TS, Junick L, et al. Radiation-Induced Changes in Normal-Appearing White Matter in Patients With Cerebral Tumors: A Diffusion Tensor Imaging Study. Int J Radiat Oncol Biol Phys 2008;70:1002–10. doi:10.1016/j.ijrobp.2007.08.020.

[20] Wang S, Wu EX, Qiu D, Leung LHT, Lau HFH-F, Khong P-LPL. Longitudinal Diffusion Tensor Magnetic Resonance Imaging Study of Radiation-Induced White Matter Damage in a Rat Model. Cancer Res 2009;69:1190–8. doi:10.1158/0008-5472.CAN-08-2661.

[21] Kerchner GA, Racine CA, Hale S, Wilheim R, Laluz V, Miller BL, et al. Cognitive Processing Speed in Older Adults: Relationship with White Matter Integrity. PLoS One 2012;7. doi:10.1371/journal.pone.0050425.

[22] Greene-Schloesser D, Robbins ME, Peiffer AM, Shaw EG, Wheeler KT, Chan MD. Radiation-induced brain injury: A review. Front Oncol 2012;2:1–18. doi:10.3389/fonc.2012.00073.

[23] Makale MT, McDonald CR, Hattangadi-Gluth JA, Kesari S. Mechanisms of radiotherapy-associated cognitive disability in patients with brain tumours. Nat Rev Neurol 2016;13:52–64. doi:10.1038/nrneurol.2016.185.

[24] Balentova S, Adamkov M. Molecular, cellular and functional effects of radiation-induced brain injury: A review. Int J Mol Sci 2015;16:27796–815. doi:10.3390/ijms161126068.

[25] Johannesen TB, Lien HH, Hole KH, Lote K, Johannesen TB, Henrik H, et al. Radiological and clinical assessment of long-term brain tumour survivors after radiotherapy. Radiother Oncol 2003;69:169–76. doi:10.1016/S0167-8140(03)00192-0.

[26] Meyers CA, Brown PD. Role and relevance of neurocognitive assessment in clinical trials of patients with CNS tumors. J Clin Oncol 2006;24:1305–9. doi:10.1200/JCO.2005.04.6086.

[27] Minniti G, De Sanctis V, Muni R, Filippone F, Bozzao A, Valeriani M, et al. Radiotherapy plus concomitant and adjuvant temozolomide for glioblastoma in elderly patients. J Neurooncol 2008;88:97–103. doi:10.1007/s11060-008-9538-0.

[28] Krex D, Klink B, Hartmann C, Von Deimling A, Pietsch T, Simon M, et al. Long-term survival with glioblastoma multiforme. Brain 2007;130:2596–606. doi:10.1093/brain/awm204.

[29] Chapman CH, Nagesh V, Sundgren PC, Buchtel H, Chenevert TL, Junck L, et al. Diffusion tensor imaging of normal-appearing white matter as biomarker for radiation-induced late delayed cognitive decline. Int J Radiat Oncol Biol Phys 2012;82:2033–40. doi:10.1016/j.ijrobp.2011.01.068.

[30] Barendsen GW. Dose fractionation, dose rate and iso-effect relationships for normal tissue responses. Int J Radiat Oncol Biol Phys 1982;8:1981–97. doi:10.1016/0360-3016(82)90459-X.

[31] Leemans A, Jeurissen B, Sijbers J, Jones DK, Jeruissen B, Sijbers J, et al. ExploreDTI: a graphical toolbox for processing, analyzing, and visualizing diffusion MR data. Proc Int Soc Magn Reson Med 2009;17:3537. doi:10.1093/occmed/kqr069.

[32] Klein S, Staring M, Murphy K, Viergever MA, Pluim JPW. Elastix: A toolbox for intensity-based medical image registration. IEEE Trans Med Imaging 2010;29:196–205. doi:10.1109/TMI.2009.2035616.

[33] Leemans A, Jones DK. The B-matrix must be rotated when correcting for subject motion in DTI data. Magn Reson Med 2009;61:1336–49. doi:10.1002/mrm.21890.

[34] Tax CMW, Otte WM, Viergever MA, Dijkhuizen RM, Leemans A. REKINDLE: Robust Extraction of Kurtosis INDices with Linear Estimation. Magn Reson Med 2015;73:794–808. doi:10.1002/mrm.25165.

[35] Kersbergen KJ, Leemans A, Groenendaal F, van der Aa NE, Viergever MA, de Vries LS, et al. Microstructural brain development between 30 and 40 weeks corrected age in a longitudinal cohort of extremely preterm infants. Neuroimage 2014;103:214–24. doi:10.1016/j.neuroimage.2014.09.039.

[36] Fischl B. FreeSurfer. Neuroimage 2012;62:774–81. doi:10.1016/j.neuroimage.2012.01.021.

[37] Vos SB, Jones DK, Viergever MA, Leemans A. Partial volume effect as a hidden covariate in DTI analyses. Neuroimage 2011;55:1566–76. doi:10.1016/j.neuroimage.2011.01.048.

[38] Jenkinson M, Beckmann CF, Behrens TEJ, Woolrich MW, Smith SM. Fsl. Neuroimage 2012;62:782–90. doi:10.1016/j.neuroimage.2011.09.015.

[39] Winkler AM, Ridgway GR, Webster MA, Smith SM, Nichols TE. Permutation inference for the general linear model. Neuroimage 2014;92:381–97. doi:10.1016/j.neuroimage.2014.01.060.

[40] Nichols T, Holmes A. Nonparametric Permutation Tests for Functional Neuroimaging. Hum Brain Funct Second Ed 2003;25:887–910. doi:10.1016/B978-012264841-0/50048-2.

[41] Holmes AP, Blair RC, Watson &NA; G, Ford I, Watson JDGG, Ford I, et al. Nonparametric Analysis of Statistic Images from Functional Mapping Experiments. J Cereb Blood Flow Metab 1996;16:7–22. doi:10.1097/00004647-199601000-00002.

[42] Eklund A, Nichols TE, Knutsson H. Cluster failure: Why fMRI inferences for spatial extent have inflated false-positive rates. Proc Natl Acad Sci 2016;113:7900–5. doi:10.1073/pnas.1602413113.

[43] Winkler AM, Ridgway GR, Douaud G, Nichols TE, Smith SM. Faster permutation inference in brain imaging. Neuroimage 2016;141:502–16. doi:10.1016/j.neuroimage.2016.05.068.

[44] Wasserstein RL, Lazar NA. The ASA’s Statement on p-Values: Context, Process, and Purpose. Am Stat 2016;70:129–33. doi:10.1080/00031305.2016.1154108.

[45] Okoukoni C, McTyre ER, Ayala Peacock DN, Peiffer AM, Strowd R, Cramer C, et al. Hippocampal dose volume histogram predicts Hopkins Verbal Learning Test scores after brain irradiation. Adv Radiat Oncol 2017;2:624–9. doi:10.1016/j.adro.2017.08.013.

[46] Devanand DP, Pradhaban G, Liu X, Khandji A, De Santi S, Segal S, et al. Hippocampal and entorhinal atrophy in mild cognitive impairment: Prediction of Alzheimer disease. Neurology 2007. doi:10.1212/01.wnl.0000256697.20968.d7.

[47] Jack CR, Petersen RC, Xu YC, O’Brien PC, Smith GE, Ivnik RJ, et al. Prediction of AD with MRI-based hippocampal volume in mild cognitive impairment. Neurology 1999;52:1397–403.

[48] Du AT, Schuff N, Amend D, Laakso MP, Hsu YY, Jagust WJ, et al. Magnetic resonance imaging of the entorhinal cortex and hippocampus in mild cognitive impairment and Alzheimer’s disease. J Neurol Neurosurg Psychiatry 2001;71:441–7. doi:10.1136/jnnp.71.4.441.

[49] Shi F, Liu B, Zhou Y, Yu C, Jiang T. Hippocampal volume and asymmetry in mild cognitive impairment and Alzheimer’s disease: Meta-analyses of MRI studies. Hippocampus 2009;19:1055–64. doi:10.1002/hipo.20573.

[50] Martin T, Grimm J, Mcintyre R, Anderson-Keightly H, Kleinberg LR, Hales RK, et al. A prospective evaluation of hippocampal radiation dose volume effects and memory deficits following cranial irradiation. Radiother Oncol 2017;125:234–40. doi:10.1016/j.radonc.2017.09.035.

[51] Nolen SC, Lee B, Shantharam S, Yu HJ, Su L, Billimek J, et al. The effects of sequential treatments on hippocampal volumes in malignant glioma patients. J Neurooncol 2016;129:433–41. doi:10.1007/s11060-016-2188-8.

[52] Blomstrand M, Kalm M, Grandér R, Björk-Eriksson T, Blomgren K. Different reactions to irradiation in the juvenile and adult hippocampus. Int J Radiat Biol 2014;90:807–15. doi:10.3109/09553002.2014.942015.

[53] Hong AM, Hallock H, Valenzuela M, Lo S, Paton E, Ng D, et al. Hippocampal Avoidance Whole Brain Radiation Therapy is Associated with Preservation of Hippocampal Volume at Six Months: A Case Series. Neuro-Oncology Open Access 2017;2:1–4. doi:10.21767/2572-0376.100017.

[54] Vos SB, Jones DK, Viergever MA, Leemans A. Partial volume effect as a hidden covariate in tractography based analyses of fractional anisotropy: Does size matter? Proc Int Soc Magn Reson Med 2010:2010.

[55] Connor M, Karunamuni R, McDonald C, Seibert T, White N, Moiseenko V, et al. Regional susceptibility to dose-dependent white matter damage after brain radiotherapy. Radiother Oncol 2017;123:209–17. doi:10.1016/j.radonc.2017.04.006.

[56] Haris M, Kumar S, Raj MK, Das KJM, Sapru S, Behari S, et al. Serial diffusion tensor imaging to characterize radiation-induced changes in normal-appearing white matter following radiotherapy in patients with adult low-grade gliomas. Radiat Med - Med Imaging Radiat Oncol 2008. doi:10.1007/s11604-007-0209-4.

[57] Khong PL, Kwong DLW, Chan GCF, Sham JST, Chan FL, Ooi GC. Diffusion-tensor imaging for the detection and quantification of treatment-induced white matter injury in children with medulloblastoma: A pilot study. Am J Neuroradiol 2003.

[58] Welzel T, Niethammer A, Mende U, Heiland S, Wenz F, Debus J, et al. Diffusion tensor imaging screening of radiation-induced changes in the white matter after prophylactic cranial irradiation of patients with small cell lung cancer: First results of a prospective study. Am J Neuroradiol 2008. doi:10.3174/ajnr.A0797.

[59] Ravn S, Holmberg M, Sorensen P, Frokjær JB, Carl J. Differences in supratentorial white matter diffusion after radiotherapy-new biomarker of normal brain tissue damage? Acta Oncol (Madr) 2013;52:1314–9. doi:10.3109/0284186X.2013.812797.

[60] Budde MD, Janes L, Gold E, Turtzo LC, Frank JA. The contribution of gliosis to diffusion tensor anisotropy and tractography following traumatic brain injury: Validation in the rat using Fourier analysis of stained tissue sections. Brain 2011;134:2248–60. doi:10.1093/brain/awr161.

[61] Douaud G, Jbabdi S, Behrens TEJ, Menke RA, Gass A, Monsch AU, et al. DTI measures in crossing-fibre areas: Increased diffusion anisotropy reveals early white matter alteration in MCI and mild Alzheimer’s disease. Neuroimage 2011;55:880–90. doi:10.1016/j.neuroimage.2010.12.008.

[62] Mac Donald CL, Dikranian K, Bayly P, Holtzman D, Brody D. Diffusion Tensor Imaging Reliably Detects Experimental Traumatic Axonal Injury and Indicates Approximate Time of Injury. J Neurosci 2007;27:11869–76. doi:10.1523/jneurosci.3647-07.2007.

[63] Walker AJ, Ruzevick J, Malayeri AA, Rigamonti D, Lim M, Redmond KJ, et al. Postradiation imaging changes in the CNS: How can we differentiate between treatment effect and disease progression? Futur Oncol 2014;10:1277–97. doi:10.2217/fon.13.271.

[64] Pasternak O, Kubicki M, Shenton ME. In vivo imaging of neuroinflammation in schizophrenia. Schizophr Res 2016;173:200–12. doi:10.1016/j.schres.2015.05.034.

[65] Trivedi R, Khan AR, Rana P, Haridas S, Hemanth Kumar BS, Manda K, et al. Radiation-induced early changes in the brain and behavior: Serial diffusion tensor imaging and behavioral evaluation after graded doses of radiation. J Neurosci Res 2012;90:2009–19. doi:10.1002/jnr.23073.

[66] Larjavaara S, Mäntylä R, Salminen T, Haapasalo H, Raitanen J, Jääskeläinen J, et al. Incidence of gliomas by anatomic location. Neuro Oncol 2007;9:319–25. doi:10.1215/15228517-2007-016.

[67] Tang Q, Lian Y, Yu J, Wang Y, Shi Z, Chen L. Anatomic mapping of molecular subtypes in diffuse glioma. BMC Neurol 2017;17:183. doi:10.1186/s12883-017-0961-8.

[68] Wijnenga MMJ, van der Voort SR, French PJ, Klein S, Dubbink HJ, Dinjens WNM, et al. Differences in spatial distribution between WHO 2016 low-grade glioma molecular subgroups. Neuro-Oncology Adv 2019. doi:10.1093/noajnl/vdz001.

[69] Nagtegaal SHJ, David S, van der Boog ATJ, Leemans A, Verhoeff JJC. Changes in cortical thickness and volume after cranial radiation treatment: A systematic review. Radiother Oncol 2019;135:33–42. doi:10.1016/j.radonc.2019.02.013.

[70] Huynh-Le M-P, Karunamuni R, Moiseenko V, Farid N, McDonald CR, Hattangadi-Gluth JA, et al. Dose-dependent atrophy of the amygdala after radiotherapy. Radiother Oncol 2019;136:44–9. doi:10.1016/j.radonc.2019.03.024.

[71] Nagtegaal S, David S, Mesri H, Philippens M, Leemans A, Verhoeff J. Identifying No Fly Zones to prevent long-term thinning of the cerebral cortex in glioma after RT. Radiother Oncol 2019;133:S82–3. doi:10.1016/s0167-8140(19)30587-0.

[72] Nobis L, Manohar SG, Smith SM, Alfaro- F, Jenkinson M, Mackay CE, et al. Hippocampal volume across age: Nomograms derived from over 19,700 people in UK Biobank. NeuroImage Clin 2019;23:1–47. doi:10.1101/562678.

[73] Ness KK, Krull KR, Jones KE, Mulrooney DA, Armstrong GT, Green DM, et al. Physiologic frailty as a sign of accelerated aging among adult survivors of childhood cancer: A report from the st jude lifetime cohort study. J Clin Oncol 2013;31:4496–503. doi:10.1200/JCO.2013.52.2268.

[74] Mandelblatt JS, Hurria A, McDonald BC, Saykin AJ, Stern RA, Vanmeter JW, et al. Cognitive effects of cancer and its treatments at the intersection of aging: What do we know; What do we need to know? Semin Oncol 2013;40:709–25. doi:10.1053/j.seminoncol.2013.09.006.

[75] Seibert TM, Karunamuni R, Bartsch H, Kaifi S, Krishnan AP, Dalia Y, et al. Radiation Dose– Dependent Hippocampal Atrophy Detected With Longitudinal Volumetric Magnetic Resonance Imaging. Int J Radiat Oncol Biol Phys 2017;97:263–9. doi:10.1016/j.ijrobp.2016.10.035.

[76] Fjell AM, Walhovd KB, Westlye LT, Østby Y, Tamnes CK, Jernigan TL, et al. When does brain aging accelerate? Dangers of quadratic fits in cross-sectional studies. Neuroimage 2010;50:1376–83. doi:10.1016/j.neuroimage.2010.01.061.

[77] Jeurissen B, Leemans A, Tournier JD, Jones DK, Sijbers J. Investigating the prevalence of complex fiber configurations in white matter tissue with diffusion magnetic resonance imaging. Hum Brain Mapp 2013;34:2747–66. doi:10.1002/hbm.22099.

[78] Vos SB, Jones DK, Jeurissen B, Viergever MA, Leemans A. The influence of complex white matter architecture on the mean diffusivity in diffusion tensor MRI of the human brain. Neuroimage 2012;59:2208–16. doi:10.1016/j.neuroimage.2011.09.086.

[79] David S, Heemskerk AM, Corrivetti F, Thiebaut de Schotten M, Sarubbo S, Corsini F, et al. The Superoanterior Fasciculus (SAF): A Novel White Matter Pathway in the Human Brain? Front Neuroanat 2019;13:1–18. doi:10.3389/fnana.2019.00024.

[80] Mesri HY, David S, Viergever M, Leemans A. Investigating the performance of Diffusional Kurtosis Imaging for group-wise analyses: A study from the Human Connectome Project. Proc. Int. Soc. Magn. Reson. Med., Paris: 2018, p. 3097.

[81] Umesh Rudrapatna S, Wieloch T, Beirup K, Ruscher K, Mol W, Yanev P, et al. Can diffusion kurtosis imaging improve the sensitivity and specificity of detecting microstructural alterations in brain tissue chronically after experimental stroke? Comparisons with diffusion tensor imaging and histology. Neuroimage 2014;97:363–73. doi:10.1016/j.neuroimage.2014.04.013.

[82] Szczepankiewicz F, Lätt J, Wirestam R, Leemans A, Sundgren P, van Westen D, et al. Variability in diffusion kurtosis imaging: Impact on study design, statistical power and interpretation. Neuroimage 2013;76:145–54. doi:10.1016/j.neuroimage.2013.02.078.

[83] Sinke MRT, Otte WM, Christiaens D, Schmitt O, Leemans A, van der Toorn A, et al. Diffusion MRI-based cortical connectome reconstruction: dependency on tractography procedures and neuroanatomical characteristics. Brain Struct Funct 2018;223:2269–85. doi:10.1007/s00429-018-1628-y.

[84] Tournier J-D, Mori S, Leemans A. Diffusion tensor imaging and beyond. Magn Reson Med 2011;65:1532–56. doi:10.1002/mrm.22924.

[85] Dell’Acqua F, Scifo P, Rizzo G, Catani M, Simmons A, Scotti G, et al. A modified damped Richardson-Lucy algorithm to reduce isotropic background effects in spherical deconvolution. Neuroimage 2010;49:1446–58. doi:10.1016/j.neuroimage.2009.09.033.

[86] Philippens M, García-Álvarez R. Functional MR Imaging. MRI Radiother. Planning, Deliv. Response Assess., Cham: Springer International Publishing; 2019, p. 73–94. doi:10.1007/978-3-030-14442-5_5.

